# Targeting multiphosphorylated tau: technical and clinical validation of a new Simoa® assay for CSF and plasma detection of tau simultaneously phosphorylated at T181 and T231

**DOI:** 10.1101/2023.06.08.23291128

**Authors:** Anna Lidia Wojdała, Giovanni Bellomo, Lorenzo Gaetani, Dandan Shan, Lucilla Parnetti, Davide Chiasserini

**Author notes:** Correspondence to: Davide Chiasserini, PhD, Section of Physiology and Biochemistry, Department of Medicine and Surgery, University of Perugia, Perugia, 06132, Italy.

## Abstract

**Background and Objectives:** Different forms of phosphorylated tau (p-tau) have shown high potential as Alzheimer’s Disease (AD) biomarkers in both cerebrospinal fluid (CSF) and plasma. Hence, we hypothesized that tau peptides showing concomitant phosphorylation at two different sites may provide an increased diagnostic value. We therefore developed and validated a new Simoa® immunoassay detecting tau simultaneously phosphorylated at T181 and T231 (C231D181) in cerebrospinal fluid (CSF) and plasma.

**Methods:** Technical validation of the C231D181 Simoa® assay included standard curve development, assessment of antibodies cross-reactivity, dilutional linearity, sensitivity, as well as intra- and inter-assay precision. Subsequently, we measured CSF C231D181, p-tau181, and p-tau231 in two cohorts: discovery (MCI-AD n=21, AD dementia n=19, CTRL n=15) and validation (preclinical AD n=19, MCI-AD n=20, AD dementia n=16, frontotemporal dementia n=39, CTRL n=24). Additionally, in the discovery cohort, C231D181, p-tau181, and p-tau231 levels were measured in matched plasma samples.

**Results:** Specificity of the assay was assessed using a synthetic peptide simultaneously phosphorylated at T181 and T231, while cross-reactivity was excluded with a mix of single-site phosphorylated peptides (T181 or T231). Both in discovery and validation cohorts, CSF C231D181, p-tau181, and p-tau231 levels were significantly elevated in all AD groups vs. CTRL. As assessed in discovery cohort, plasma p-tau231 and p-tau181 levels enabled effective discrimination of AD continuum groups from CTRL (AUC plasma p-tau231: CTRL vs. MCI-AD=0.925, CTRL vs. AD-dem=0.947; AUC plasma p-tau181: CTRL vs. MCI-AD=0.877, CTRL vs. AD-dem=0.943) while plasma C231D181 did not change among clinical groups.

**Discussion:** A new ultrasensitive immunoassay detecting tau simultaneously phosphorylated at T181 and T231 was developed and validated. While we found this phosphorylated tau form to be significantly elevated across the AD continuum in CSF, in plasma it did not show changes among the diagnostic groups. The differences between CSF and plasma suggest matrix-specific protein processing. Our findings support evidence for qualitative and quantitative importance of tau phosphorylation across AD continuum and warrant further investigation, including assessment of tau simultaneously phosphorylated at multiple sites.

## 1. Background

Alzheimer’s disease (AD) is a progressive neurodegenerative disease which affects a constantly growing part of the population, accounting for 60% to 80% of all dementia cases [1]. The most characteristic hallmarks of the disease include accumulation of extracellular amyloid plaques, intraneuronal neurofibrillary tangles, and brain atrophy caused by neuronal and synaptic/axonal degeneration [2]. It is currently well established that these changes start years before the onset of clinical symptoms [3,4]. In the fast-developing field of fluid AD biomarkers, considerable efforts have been made to discover and establish tools enabling detection of such alterations at the early stage of the disease, before neuronal damage reaches an advanced and not reversible phase. Thanks to its direct connection with central nervous system (CNS), cerebrospinal fluid (CSF) is considered as a primary matrix of interest for detection of AD biomarkers. The main biomarkers already implemented in the AD diagnostic process are beta-amyloid peptides (Aβ42, Aβ40, and Aβ42/Aβ40 ratio), total tau (t-tau), and different forms of phosphorylated tau (primarily, p-tau181, and to some extent, p-tau231 and p-tau217) [5,6]. Thanks to development of ultrasensitive technologies, extending range of measurable protein concentrations, last decade resulted in a rain of studies exploring diagnostic potential of AD biomarkers measured in plasma. Likewise in CSF, phosphorylated forms of tau (namely – p-tau181, p-tau217, p-tau231) measured in plasma have been shown to provide a high diagnostic value across different stages of the AD continuum [7–14]. Additionally, phosphorylated tau forms like p-tau217 and p-tau231 have been reported to be associated with disease stage, showing an increase at very early phases of the disease [15–17]. However, diverse tau posttranslational modifications (PTMs) have been identified at more than 50 sites [18] and their importance, both for physiological and pathological stage, is not fully understood yet. Tau phosphorylation regulates microtubules binding, hence impacts axon outgrowth and axonal transport - processes known to be impaired in AD pathology [19]. Recent studies raised the question whether tau hyperphosphorylation might be a physiological process which intensifies in the course of AD progression [18,20,21].

To investigate AD biomarker potential of multiphosphorylated tau in human biofluids, we designed a new sandwich immunoassay exclusively detecting protein simultaneously phosphorylated at two distinct sites (T181, T231). Subsequently, we analyzed levels of tau simultaneously phosphorylated at T181 and T231 (C231D181) as well as levels of p-tau181 and p-tau231 (single-site phosphorylation) in matched CSF and plasma samples originating from two well-characterized cohorts of patients across the AD continuum (discovery cohort n=55, validation cohort, n=118). AD patients were compared with control subjects without cognitive decline (in both cohorts) and with frontotemporal dementia (FTD) patients (in the validation cohort). Noteworthy, besides mild cognitive impairment due to AD (MCI-AD) and AD at the stage of dementia (AD-dem, both cohorts), the validation cohort also included patients at very early, asymptomatic disease stage, i.e., functionally independent patients with no cognitive deficits but with a positive CSF AD biomarker profile (pre-AD). The results show that our ultra-sensitive assay is capable of detecting tau peptides with double phosphorylation at T181 and T231 in both CSF and plasma and can be used for screening larger cohorts of AD continuum patients.

## 2. Methods

### 2.1. Study participants

Two patients’ cohorts (discovery cohort n=55, validation cohort n=118) from the Neurology Clinic, S. Maria della Misericordia Hospital (Perugia, Italy) were retrospectively and consecutively enrolled in the study. All patients underwent a standardized assessment including medical history, physical and neurological examination, laboratory tests, and neuropsychological evaluation including, among screening tests, Mini-Mental State Examination (MMSE). Brain imaging (computed tomography or MRI) or ^18^Fluoro-2-deoxyglucose positron emission tomography (FDG-PET), were also performed in selected cases, according to clinical suspicion. In all cases, the diagnostic process was supported by the analysis of core CSF AD biomarkers (Aβ42, Aβ40, Aβ42/Aβ40 ratio, t-tau, p-tau181) measured with the Lumipulse® G600II (Fujirebio, Japan). Following principles of A/T/N system which defines three core features of the disease – brain amyloidosis, tauopathy, and neurodegeneration occurring over the disease progression [22], the patients were therefore classified as A+/A-, T+/T-, N+/N-according to the cut-off values (Aβ42/Aβ40=0.072, 95% CI 0.07-0.074; t-tau=50, 95% CI 46.2-52.3; p-tau181=393, 95% CI 359-396) [23]. Patients within the AD continuum were all defined by an A+/T+ CSF profile, according to NIA-AA criteria [22]. The discovery cohort included patients with AD at dementia stage (AD-dem, n=19) and patients with mild cognitive impairment due to AD (MCI-AD, n=21). The AD continuum group in the validation cohort included AD-dem (n=35), MCI-AD (n=41), and preclinical Alzheimer’s disease patients (pre-AD, n=19). The pre-AD patients were subjects with subjective cognitive complaints not confirmed by neuropsychological assessment but with A+/T+ CSF profile. In the validation cohort, we also included a subgroup of patients (n=39) affected by FTD whose diagnosis was supported by FDG-PET [24,25], as a partial overlap between the clinical picture of AD and FTD patients exists. Each cohort included a control group (CTRL, discovery cohort n=15, validation cohort n=24), composed of patients diagnosed with minor neurological diseases, other than inflammatory or degenerative disease of the CNS or of the peripheral nervous system. All patients categorized as CTRL were negative for classical AD CSF biomarkers (A-/T-/N-profile). Demographic and clinical data of each patient were confidentially stored in an online electronic database.

Age, gender, years of education, and MMSE scores for each clinical group are reported in the Table 1. A/T/N profile data are reported in the Supplementary Table 1.

**Table 1.**
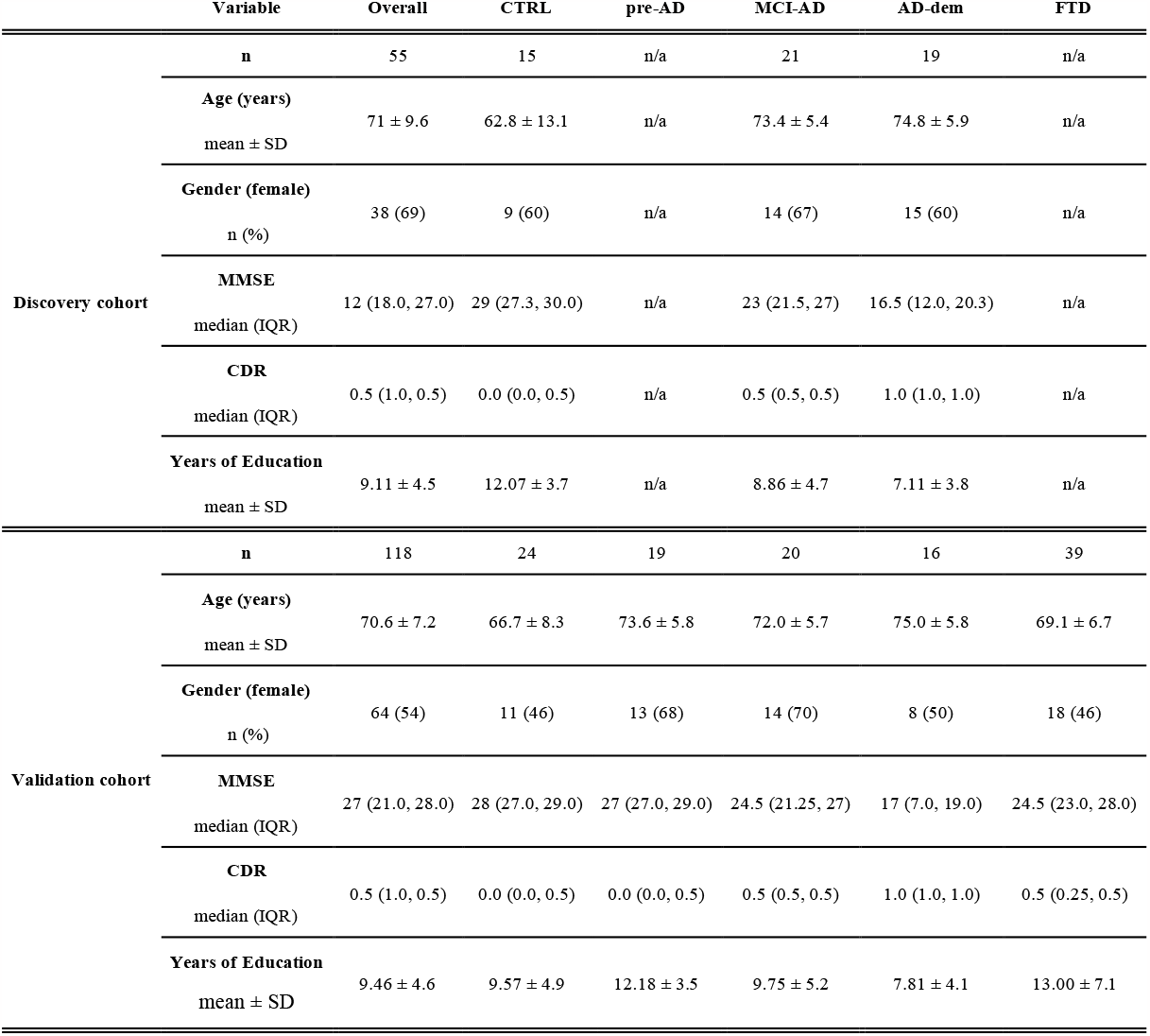
Demographic and clinical profile of the discovery and validation cohorts. MMSE: Mini-Mental State Examination; CDR: Clinical Dementia Rating; IQR: interquartile range; SD: standard deviation. AD: Alzheimer’s disease, pre-AD: preclinical AD, MCI-AD: mild cognitive impairment due to AD, AD-dem: AD at dementia stage, FTD: frontotemporal dementia, CTRL: control group.

All the procedures were performed in accordance with Declaration of Helsinki and International Conference on Harmonization guidelines for Good Clinical Practice. All the patients and/or their legal representatives gave informed written consent for the lumbar puncture and the inclusion in the study that was approved by the local Ethics Committee (Comitato Etico Aziende Sanitarie Regione Umbria 19369/AV and 20942/21/OV).

### 2.2. Collection of human samples

CSF and plasma samples were collected between 2012 and 2021, following international guidelines and the same standard operating procedures throughout the study [26,27]. LP was performed between 8:00 and 10:00 a.m. CSF was collected into sterile polypropylene tubes and centrifuged for 10 min at 2000 × g at room temperature (RT). At the same time, plasma was collected into sterile polypropylene tubes containing EDTA as the anticoagulant and centrifuged for 10 min at 2000 × g (RT). Once processed, CSF and plasma samples were stored in 0.5 mL tubes (72.730.007, Sarstedt, Germany) and immediately frozen at −80°C pending analysis.

### 2.3. Development of Simoa® assay for detection of multiphosphorylated tau in CSF and plasma

A 2-step protocol was used to develop Simoa® sandwich immunoassay for CSF and plasma detection of tau simultaneously phosphorylated at T181 and T231. All the measurements were run on the HD-X™ instrument (Quanterix, USA).

Single Molecule Array, acronymized as Simoa®, is a bead-based immunoenzymatic technology enabling detection of proteins at sub-femtomolar levels [28]. Briefly, sandwich immunocomplexes are formed by incubation of a protein target with capture antibody-coated paramagnetic beads and enzyme-labelled (streptavidin beta-galactosidase, SBG) detector antibody. Immunocomplexes are resuspended in a solution of the SBG substrate - resorufin-beta-D-galactopyranoside (RGP) and then transferred to microarray Simoa® discs. Over 200,000 thousands of 50-femtoliter wells distributed on each array are big enough to host only one bead per time, hence immunocomplexes are effectively separated. Once the bead loading is completed, an oily layer is spread, sealing beads in wells. As the enzyme-bound complexes and RGP substrate are mixed together, an enzymatic reaction takes place and results in generation of resorufin, a fluorescent reaction product. The increase of fluorescent signal for single beads is recorded and converted into Simoa® unit of measurement - Average Enzymes per Bead (AEB). In presence of the proper calibrator AEB values are further interpolated into pg/mL.

In the C231D181 assay, paramagnetic beads were conjugated with monoclonal antibody specific for tau phosphorylated at T231. Monoclonal antibody targeting tau phosphorylated at T181 was labelled with SBG enzyme and used as a detector antibody. Capture (p-tau231) antibody used in C231D181 assay was the antibody used as a capture antibody in p-tau231 Advantage Kit (item 102292 Quanterix, USA). Detector (p-tau181) antibody used in C231D181 assay corresponded to the antibody used as a capture antibody in p-tau181 Advantage V2 Kit (item 103714 Quanterix, USA). 4-fold dilution in N4PE CSF Sample Diluent (Quanterix, USA) was established as the default CSF and plasma sample dilution for C231D181 assay. A synthetic peptide, specifically phosphorylated at T181 and T231 (N-term→C-term sequence: IPAKTPPAPKT(PO_3_H_2_)PPSSGEPPKREPKKVAVVRT (PO_3_H_2_)PPKSPSSAK; Anaspec, USA) was used as the assay calibrator. Peptide standard dilutions were prepared with use of the N4PE CSF Sample Diluent (Quanterix, USA).

The C231D181 assay underwent a technical validation. This consisted in the estimation of the intermediate precision (inter-assay reproducibility within the laboratory), repeatability (intra-assay), dilution linearity, and sensitivity (Lower Limit of Quantification, LLOQ; Upper Limit of Quantification, ULOQ). Range of the standard curve was established, and the intra- and inter-assay variabilities were assessed based on coefficient of variation (CV) of the replicates. Dilution linearity, assessed on two independent instruments, was assessed for the sample spiked with the peptide standard. Mix of single-site phosphorylated tau peptides (phosphorylation at T181 or at T231) was used to exclude antibodies cross-reactivity. Four parameter logistic (4PL) curve model with 1/y weighting was used for curve fitting.

As reference, CSF and plasma levels of tau phosphorylated at threonine 181 (p-tau181) and tau phosphorylated at threonine 231 (p-tau231) were measured with in-house manufactured Simoa® assays, utilizing the same antibodies, standards, and buffers as the commercially available kits offered by Quanterix i.e., p-tau181 Advantage V2 Kit (item 103714) and p-tau231 Advantage Kit (item 102292). Appropriate calibrators and controls included in each kit were run together with samples. After each run, it was controlled whether the measured concentration of controls fit into the lot-specific range. Samples were anonymized and the researcher completing measurements was blinded to patients’ clinical profile.

### 2.4. Statistical Analysis

Continuous variables are represented as the median ± interquartile range (IQR). Statistical analysis was performed with the use of the GraphPad Prism software version 9.5.0 (USA) and R software version 4.2.2. [29]. D’Agostino-Pearson omnibus K2 test was applied to assess data normality. Non-parametric Kruskal-Wallis test was applied for multiple group comparisons, followed by Dunn’s test for multiple comparisons correction. Correlation matrix was assessed by non-parametric Spearman correlation (p-value two-tailed, 95% confidence intervals (CI)). Receiver operating characteristic curve (ROC) (95% CI, Wilson/Brown method) analysis was applied to calculate the accuracy of each assay to discriminate between control group and different AD continuum stages. A *p* value ≤ 0.05 was considered statistically significant for all the analyses.

## 3. Results

### 3.1. Technical validation of the C231D181 assay

The 8-point standard curve of the C231D181 assay included peptide standard dilutions (1:2) ranging from 80 pg/mL to 1.25 pg/m and a blank, devoted of peptide standard. LLOQ and ULOQ were determined as 1.25 pg/mL and 80 pg/mL, respectively. The CV% for the inter- and intra-assay variability were, respectively, 10% and 3%. Good dilution linearity results were obtained (CV% on instrument 1: 11%, instrument 2: 6%; 1x, 2x, 4x, 8x, 16x, 32x dilutions of a spiked sample tested). Measurement of 30 pg/mL, 10 pg/mL, 3.33 pg/mL, 1.11 pg/mL, 0.370 pg/mL, 0.123 pg/mL, and 0.041 pg/mL of the single-site phosphorylated peptides mix resulted in AEB signal equal to blank (AEB approx. 0.005), hence confirmed lack of antibodies cross-reactivity. Detail information regarding technical validation can be found in the Supplementary Tables 2-6.

### 3.2. Demographic and clinical profile of the cohort

Demographics and clinical parameters of the two cohorts are reported in Table 1. In both cohorts, statistically significant differences in age were observed among selected groups (discovery: CTRL vs. MCI-AD *p* ≤ 0.01, CTRL vs. AD-dem *p* ≤ 0.001, validation: CTRL vs. pre-AD, *p* ≤ 0.05, CTRL vs. AD-dem *p* ≤ 0.01, AD-dem vs. FTD *p* ≤ 0.05; Kruskal-Wallis, followed by Dunn’s *post hoc* test). No significant gender differences were present in any of the cohorts (Chi-square test). As expected, in both cohorts baseline Mini-Mental State Examination (MMSE) scores differed among the groups (*p* ≤ 0.0001, Kruskal-Wallis). In the discovery cohort, *post hoc* analysis (Dunn’s test for multiple comparisons) revealed statistically significant decrease in MMSE scores for MCI-AD group vs. CTRL (*p* ≤ 0.01), AD-dem vs. CTRL (*p* ≤ 0.0001), and AD-dem vs. MCI-AD (*p* ≤ 0.01). In the validation cohort, statistically significant difference in MMSE scores was found for MCI-AD vs. CTRL (*p* ≤ 0.05), AD-dem vs. CTRL (*p* ≤ 0.0001), pre-AD vs. MCI-AD (*p* ≤ 0.05), pre-AD vs. AD-dem (*p* ≤ 0.05), MCI-AD vs. AD-dem (*p* ≤ 0.05), and AD-dem vs. FTD (*p* ≤ 0.05), while no difference was observed for pre-AD vs. CTRL, FTD vs. CTRL, pre-AD vs. FTD, and MCI-AD vs. FTD.

As assessed by Lumipulse® CSF measurement, all subjects within the CTRL group were characterized by A-T-N-CSF profile. The profile of pre-AD and MCI-AD patients was A+/T+. The majority of AD patients had A+T+N+ profile with an exception of two patients (1 patient in the discovery cohort and 1 patient in the validation cohort) having biomarker values within cut-off 95% CI and clinical picture clearly pointing to AD at the stage of dementia [23].

### 3.3. Correlations between CSF and plasma biomarkers

As assessed jointly in combined discovery and validation cohorts, CSF C231D181 strongly correlated with CSF levels of p-tau181 (ρ = 0.93) and p-tau231 (ρ = 0.93) (Spearman correlation analysis). In the discovery cohort, plasma C231D181 did not correlate with CSF and plasma p-tau181, p-tau231 nor with its CSF equivalent (CSF C231D181) (Fig. 1).

**Fig. 1.**
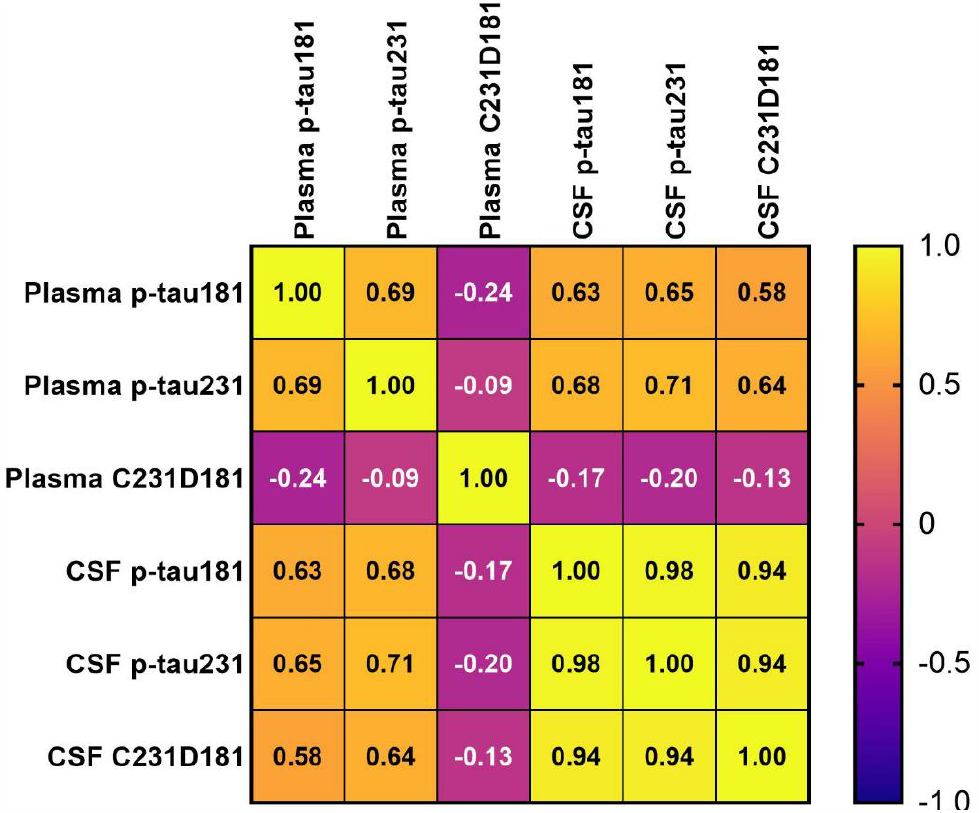
Correlations among CSF and plasma biomarkers. Heatmap presenting results of Spearman correlation analysis for CSF and plasma biomarkers assessed across the whole discovery cohort (CTRL, MCI-AD, AD-dem). The color gradient indicates the strength of correlation according to Spearman ρ.

### 3.4. Diagnostic performance in the discovery cohort

CSF p-tau181 levels were assessed in the whole cohort by Lumipulse® and used to assess T profile within A/T/N system. However, to harmonize measurements in CSF and plasma on a single platform, all the CSF samples underwent Simoa® p-tau181 assessment to be subsequently compared with related plasma p-tau181 Simoa® measurements.

In the discovery cohort, CSF p-tau181, p-tau231, and C231D181 levels were consistently increased in pre-AD, MCI-AD, and AD-dem groups while compared with controls (*p* ≤ 0.0001) (Fig. 2A). Plasma p-tau181 and p-tau231 levels were significantly increased in MCI-AD and AD-dem groups vs. CTRL (plasma p-tau181: CTRL vs. MCI-AD *p* ≤ 0.01, CTRL vs. AD-dem *p* ≤ 0.0001; plasma p-tau231: CTRL vs. MCI-AD *p* ≤ 0.001, CTRL vs. AD-dem *p* ≤ 0.0001). Plasma C231D181 levels did not significantly differ between CTRL and any of AD continuum groups (Fig. 2B).

**Figure 2.**
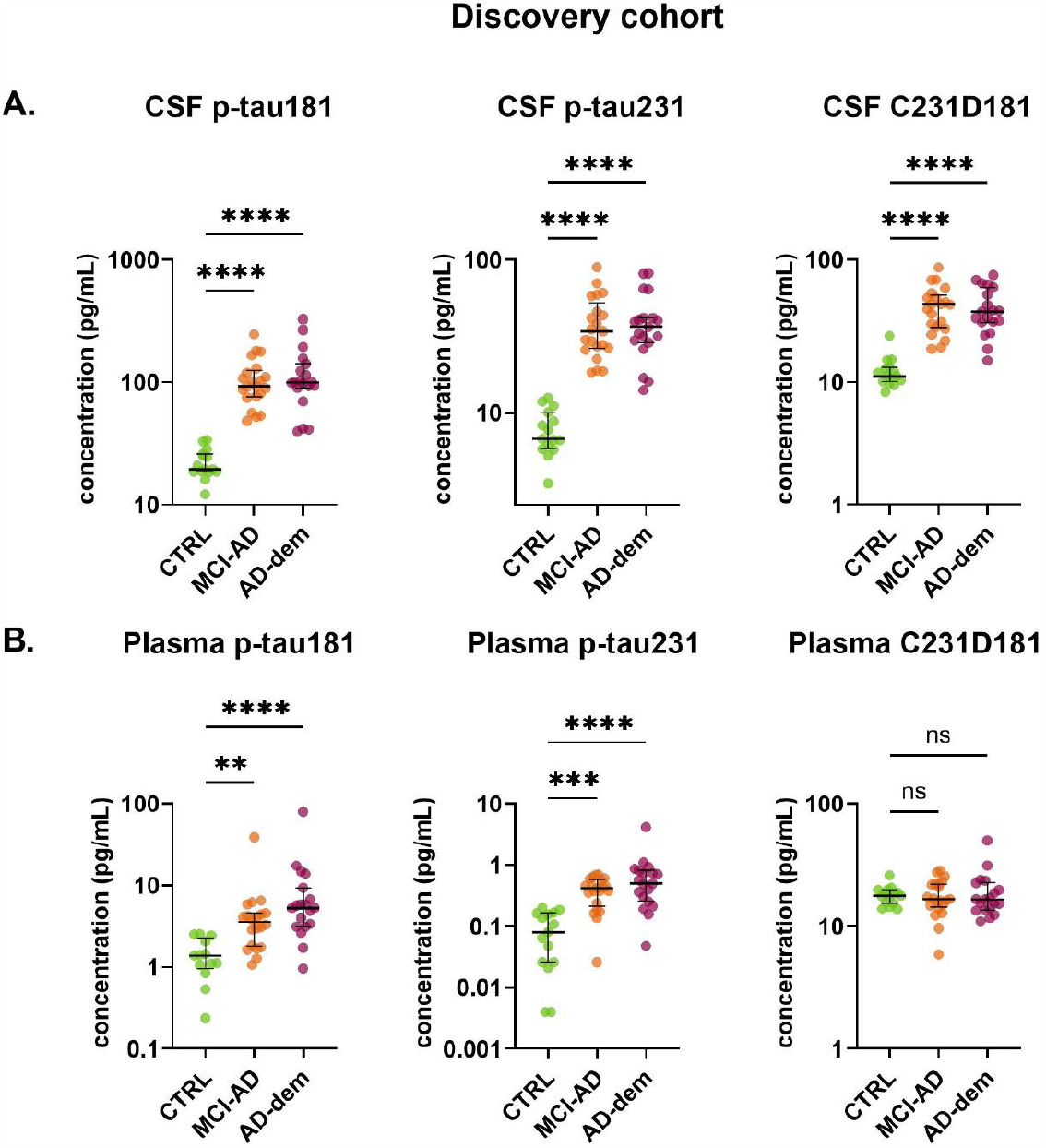
Diagnostic performance CSF and plasma C231D181 assay and reference assays (p-tau181, p-tau231) in the discovery cohort. A. CSF, B. plasma measurements. Horizontal line refers to median and error bars indicate interquartile range. Data plotted in log(10) scale. Legend: ns = *p* > 0.05, * = *p* ≤ 0.05, ** = *p* ≤ 0.01, *** = *p* ≤ 0.001, **** = *p* ≤ 0.0001, comparison vs. CTRL. AD: Alzheimer’s disease, pre-AD: preclinical AD, MCI-AD: mild cognitive impairment due to AD, AD-dem: AD at dementia stage, CTRL: control group.

ROC analysis performed for CSF p-tau181, p-tau231, and C231D181 measured in the discovery cohort, showed a high accuracy in discriminating between CTRL and AD continuum stages (AUC range 0.986-1.000; Supplementary Table 7). In plasma, p-tau231 exhibited the best ability to differentiate between CTRL and both MCI-AD and AD-dem (plasma p-tau231 CTRL vs. MCI-AD AUC = 0.925, 95% CI 0.840-1.000; CTRL vs. AD-dem AUC = 0.947, 95% CI 0.872-1.000), followed by p-tau181 (plasma p-tau181 CTRL vs. MCI-AD AUC = 0.877, 95% CI 0.761-0.993; CTRL vs. AD-dem AUC = 0.943, 95% CI 0.857-1.000) (Fig. 3). ROC analysis performed for plasma C231D181 showed low performance in discriminating CTRL vs. MCI-AD (AUC = 0.514, 95% CI 0.320-0.709) and CTRL vs. AD-dem (AUC = 0.579, 95% CI 0.383-0.775).

**Fig. 3.**
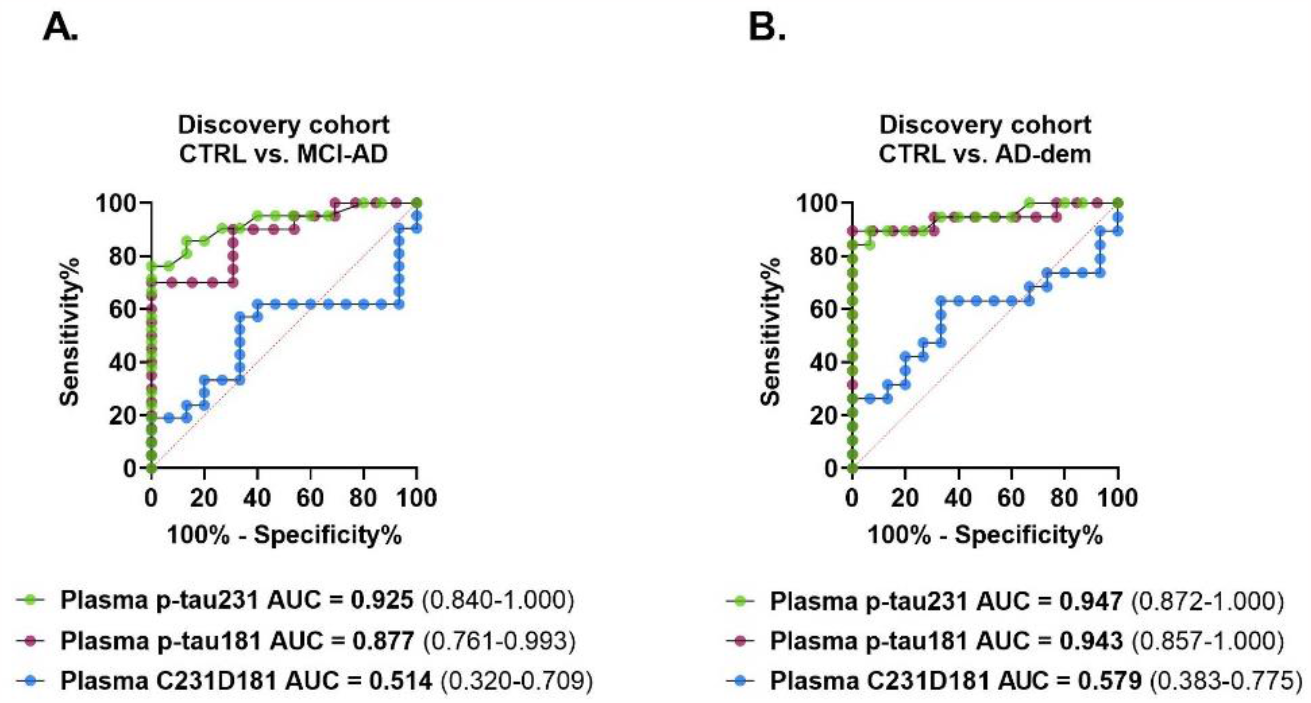
Panel of superimposed plasma p-tau181, p-tau231, and C231D181 ROC curves for discrimination of CTRL vs. MCI-AD (A.) and CTRL vs. AD-dem (B.) groups. AUC: Area Under Curve. Values in brackets represent CI 95 % (Wilson/Brown method).

### 3.5. Diagnostic performance in the validation cohort

In the validation cohort, CSF p-tau181, p-tau231, and C231D181 levels were consistently elevated in pre-AD, MCI-AD, and AD-dem groups while compared with controls (*p* ≤ 0.0001). CSF p-tau181, p-tau231, and C231D181 levels did not significantly differ between CTRL and FTD in any of the cohorts (Fig. 4).

**Figure 4.**
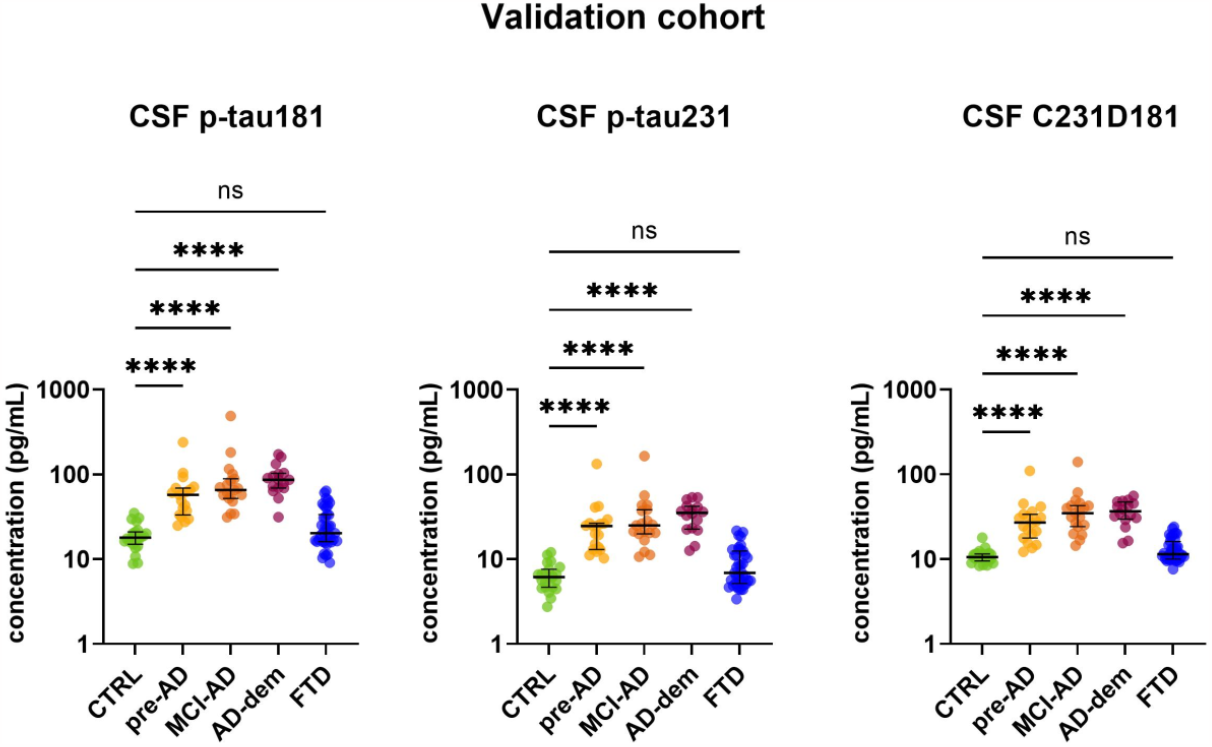
Diagnostic performance CSF C231D181 assay and reference assays in (p-tau181, p-tau231) in the validation cohort. Horizontal line refers to median and error bars indicate interquartile range. Data plotted in log(10) scale. Legend: ns = *p* > 0.05, * = *p* ≤ 0.05, ** = *p* ≤ 0.01, *** = *p* ≤ 0.001, **** = *p* ≤ 0.0001, comparison vs. CTRL. AD: Alzheimer’s disease, pre-AD: preclinical AD, MCI-AD: mild cognitive impairment due to AD, AD-dem: AD at dementia stage, FTD: frontotemporal dementia, CTRL: control group.

ROC analysis performed for CSF p-tau181, p-tau231, and C231D181 measured in the validation cohort confirmed high accuracy of all three assays in discriminating between CTRL and MCI-AD and CTRL vs. AD-dem (AUC range = 0.991-1.000; Supplementary Table 7). Also ROC analysis of CTRL vs. pre-AD resulted in high AUC values for each of the assays (CSF p-tau181 CTRL vs. pre-AD AUC = 0.970, 95% CI 0.929-1.000; CSF p-tau231 AUC = 0.991, 95% CI 0.972-1.000; CSF C231D181 AUC = 0.982, 95% CI 0.951-1.000). CSF measurement of p-tau181, p-tau231, and C231D181 allowed for an effective discrimination between AD-dem vs. FTD (AUC range = 0.968-0.978) in the validation cohort (Supplementary Table 7).

## 4. Discussion

In the current study we developed and validated C231D181 - a novel assay targeting tau simultaneously phosphorylated at T181 and T231. We enrolled a well-characterized discovery cohort of patients across the AD continuum (MCI-AD, AD-dem) and control subjects to assess the diagnostic potential of the new assay in CSF and plasma. CSF C231D181 exhibited a diagnostic value, comparable with classical assays targeting only one tau phosphorylation site (p-tau181, p-tau231). At the same time, we observed that plasma C231D181 measurement did not enable effective discrimination among clinical groups. Considering promising results obtained for CSF C231D181, we assessed its clinical performance in a larger validation cohort. Next to the clinical groups included in the discovery cohort (MCI-AD, AD-dem, CTRL), the validation cohort included pre-AD patients i.e., functionally independent patients with no cognitive deficits and concurrent presence of positive core CSF AD biomarker profile. Additionally, FTD patients were enrolled to assess the specificity of the biomarkers against a neurodegenerative disorder which clinical profile partially overlaps with AD. CSF C231D181 measurements in the validation cohort confirmed its good diagnostic potential as AD biomarker.

From the technical point of view, our work presents an innovative approach to target phosphorylated tau species in biological fluids. The C231D181 assay enables the sensitive and specific detection of a tau peptide carrying parallel phosphorylation at two different sites.

There is an ongoing debate whether increasing degree of tau phosphorylation reflects AD progression as well as what is the pattern and dynamics of tau phosphorylation characterizing AD [18,20,21]. Interestingly, Barthélemy et al. [30] observed that in the dominantly inherited AD cohort, increasing levels of t-tau and p-tau205 were associated with higher tau-PET levels, whereas trend of negative correlation was observed between p-tau217 and p-tau181 levels and tau-PET. Therefore, the features of tau phosphorylation across the AD spectrum and their relationship between brain and biofluids requires further investigation.

Current literature evidence suggests that majority of CSF tau is truncated and present as mainly N-terminal fragments with different cleavage sites (predominantly within positions 221-226) [20,31,32]. A similar fragmentation pattern has also been found in plasma, since a significant decrease of tau peptides spanning the 221-226 positions has been observed [11]. The existence of these truncations in biofluids may result in low abundance of the peptide spanning from 181 to 231 residues. However, the high sensitivity of the Simoa® technology enables its detection with our optimized protocol. The evidence for presence of peptides covering residues 181 and 231 and bearing parallel phosphorylation at the two sites was shown by Ashton and colleagues [13]. The authors applied targeted mass spectrometry methods to identify a) phosphorylation at T231 in p-tau181 immunodepleted CSF pool, b) to identify phosphorylation at T181 in p-tau231 immunodepleted CSF pool [13].

In our study, diagnostic performance of C231D181 significantly differs between CSF and plasma. An observed lack of agreement between matrices might be due to differences in each matrix environment which result in the altered tau processing [33]. Additionally, peripheral tau production may influence plasma measurements [34].

While CSF and plasma increase of p-tau181, p-tau217, and p-tau231 across AD continuum is already widely reported in the literature [8,10,13,15,35], none of these studies employes immunoenzymatic methods to assess qualitative change of biofluid tau phosphorylation over the disease course. Following previous evidence suggesting that degree of tau hyperphosphorylation reflects disease progression, it should be further considered that the pattern of multiphosphorylation as well as other post-translational modifications may specifically change depending on the e.g., disease stage or presence of co-pathology [18,20,21,30]. The presented C231D181 assay design offers high flexibility, as incorporation of different antibodies, targeting other phosphorylation sites with reported AD relevance is possible. Development of such assays and assessment of their diagnostic performance would provide an important insight into p-tau metabolism across the AD continuum.

## 5. Limitations

The main limitation of the study is lack of absolute quantification of the synthetic peptide used as a calibrator for the C231D181 assay. Hence, presented concentrations of tau simultaneously phosphorylated at T181 and T231 should not be considered as definitive. Most probably, use of absolutely quantified peptide as a standard would result in more precise values. Other limitations are related to the retrospective character of the study and the sample size, especially of the pre-AD group. However, the inclusion of a group of functionally independent patients with no objective evidence of cognitive deficits but with positive core CSF AD biomarkers is fundamental to fully understand the early dynamics of biomarker changes in biofluid across the AD continuum.

## 6. Conclusions

In conclusion, we propose a novel ultrasensitive approach to target tau simultaneously phosphorylated at T181 and T231. Our results suggest its high diagnostic potential as a CSF but not plasma AD biomarker. The differences observed between CSF and plasma suggest matrix-specific protein processing and warrant further investigation, including assessment of tau simultaneously phosphorylated at sites different than those already analyzed.

## Data Availability

The datasets used and/or analyzed during the current study are available from the corresponding author on reasonable request.

## Acknowledgements

We thank Dr. Silvia Paciotti, Dr. Alfredo Megaro, and Andrea Toja for the lab support and Dr. Elena Chipi for the scientific discussion. We also thank all the Members of the MIRIADE consortium for collaboration and scientific discussion.

