## Supplementary material for "Targeting multiphosphorylated tau: technical and clinical validation of a new Simoa® assay for CSF and plasma detection of tau simultaneously phosphorylated at T181 and T231": C231D181_supplementary_material

|  | **ATN profile, n (%)** | **Overall** | **CTRL** | **pre-AD** | **MCI-AD** | **AD-dem** | **FTD** |
| --- | --- | --- | --- | --- | --- | --- | --- |
| **Discovery cohort** | **n** | 55 | 15 | n/a | 21 | 19 | n/a |
|  | A-T-N- | 15 | 15 (100) | n/a | 0 (0) | 0 (0) | n/a |
|  | A+T-N- | 1 | 0 (0) | n/a | 0 (0) | 1 (5) | n/a |
|  | A+T+N- | 0 (0) | 0 (0) | n/a | 0 (0) | 0 (0) | n/a |
|  | A+T+N+ | 21 | 0 (0) | n/a | 21 (100) | 18 (95) | n/a |
|  | A-T-N+ | 0 (0) | 0 (0) | n/a | 0 (0) | 0 (0) | n/a |
|  | A-T+N- | 0 (0) | 0 (0) | n/a | 0 (0) | 0 (0) | n/a |
|  | A-T+N+ | 0 (0) | 0 (0) | n/a | 0 (0) | 0 (0) | n/a |
| **Validation cohort** | **n** | 118 | 24 | 19 | 20 | 16 | 39 |
|  | A-T-N- | 42 (36) | 24 (100) | 0 (0) | 0 (0) | 0 (0) | 18 (46) |
|  | A+T-N- | 5 (4) | 0 (0) | 0 (0) | 0 (0) | 0 (0) | 5 (13) |
|  | A+T+N- | 10 (8) | 0 (0) | 7 (37) | 2 (10) | 0 (0) | 1 (3) |
|  | A+T+N+ | 47 (40) | 0 (0) | 12 (63) | 18 (90) | 15 (94) | 2 (5) |
|  | A-T-N+ | 2 (2) | 0 (0) | 0 (0) | 0 (0) | 0 (0) | 2 (5) |
|  | A-T+N- | 4 (3) | 0 (0) | 0 (0) | 0 (0) | 0 (0) | 4 (10) |
|  | A-T+N+ | 8 (7) | 0 (0) | 0 (0) | 0 (0) | 1 (6) | 7 (18) |

**Supplementary Table 1. A/T/N profile of the discovery and validation cohorts.** A/T/N profile assessed by measurement of CSF Aβ42, Aβ40, Aβ42/Aβ40, t-tau, and p-tau181 by Lumipulse® G600II (Fujirebio, Japan)(1). AD: Alzheimer’s disease, pre-AD: preclinical AD, MCI-AD: mild cognitive impairment due to AD, AD-dem: AD at dementia stage, FTD: frontotemporal dementia, CTRL: control group.

| **Calibrator** | **Peptide conc. (pg/mL)** | **AEB** |
| --- | --- | --- |
| A | 0 | 0.004 |
| B | 1.25 | 0.010 |
| C | 2.5 | 0.031 |
| D | 5 | 0.109 |
| E | 10 | 0.399 |
| F | 20 | 1.452 |
| G | 40 | 5.083 |
| H | 80 | 14.713 |

**Supplementary Table 2. C231D181 assay - standard curve development.**

| **Instrument** | **Dilution** | **Conc. DF corrected (pg/mL)** | **AVG conc. (pg/mL)** | **CV%** |
| --- | --- | --- | --- | --- |
| Spiked plasma sample  Instrument 1 | 1x | 34.701 | 31.605 | 11% |
|  | 2x | 32.770 |  |  |
|  | 4x | 29.892 |  |  |
|  | 8x | 27.721 |  |  |
|  | 16x | 28.631 |  |  |
|  | 32x | 35.917 |  |  |
| Spiked plasma sample  Instrument 2 | 1x | 34.031 | 31.7354 | 6% |
|  | 2x | 34.156 |  |  |
|  | 4x | 31.459 |  |  |
|  | 8x | 29.872 |  |  |
|  | 16x | 31.243 |  |  |
|  | 32x | 29.651 |  |  |

**Supplementary Table 3. C231D181 assay - dilution linearity.**

| **Instrument 1** | **Standard** | **Peptide standard (pg/mL)** | **AEB** | **AVG AEB** | **AEB CV%** | **conc. (pg/mL)** | **AVG conc. (pg/mL)** | **Recovery %** | **CV% conc.** |
| --- | --- | --- | --- | --- | --- | --- | --- | --- | --- |
|  | C | 2.5 | 0.033 | 0.029 | 0.220 | 2.416 | 2.206 | 88% | 13% |
|  |  |  | 0.024 |  |  | 1.997 |  |  |  |
|  | **B** | **1.25** | **0.010** | **0.010** | **0.060** | **1.159** | **1.159** | **93%** | **0%** |
|  |  |  | **0.010** |  |  | **1.159** |  |  |  |
|  | B/2 | 0.625 | 0.011 | 0.008 | 0.436 | 1.233 | 1.027 | 164% | 28% |
|  |  |  | 0.006 |  |  | 0.820 |  |  |  |
|  | B/4 | 0.3125 | 0.015 | 0.009 | 0.951 | 1.499 | 0.984 | 315% | 74% |
|  |  |  | 0.003 |  |  | 0.470 |  |  |  |
| **Instrument 2** | **Standard** | **Peptide standard (pg/mL)** | **AEB** | **AVG AEB** | **AEB CV%** | **conc. (pg/mL)** | **AVG conc. (pg/mL)** | **Recovery %** | **CV% conc.** |
|  | C | 2.5 | 0.026 | 0.025 | 0.055 | 2.147 | 2.097 | 84% | 3% |
|  |  |  | 0.024 |  |  | 2.048 |  |  |  |
|  | **B** | **1.25** | **0.009** | **0.009** | **0.021** | **1.122** | **1.122** | **90%** | **0%** |
|  |  |  | **0.009** |  |  | **1.122** |  |  |  |
|  | B/2 | 0.625 | 0.003 | 0.005 | 0.440 | 0.512 | 0.733 | 117% | 42% |
|  |  |  | 0.007 |  |  | 0.953 |  |  |  |
|  | B/4 | 0.3125 | 0.006 | 0.004 | 0.645 | 0.859 | 0.604 | 193% | 60% |
|  |  |  | 0.002 |  |  | 0.349 |  |  |  |

**Supplementary Table 4. C231D181 assay - lower limit of quantification (LLOQ).**

| **Run** | **Instrument** | **conc. (pg/mL)** |
| --- | --- | --- |
| Run 1 | Instrument 2 | 33.450 |
| Run 2 | Instrument 1 | 38.279 |
| Run 3 | Instrument 2 | 39.359 |
| Run 4 | Instrument 2 | 32.149 |
| Run 5 | Instrument 2 | 36.079 |
| Run 6 | Instrument 2 | 39.810 |
| **AVG conc. (pg/mL)** | | 36.521 |
| **CV%** | | **9%** |

**Supplementary Table 5. C231D181 assay – inter-assay variability.**

| **Sample** | **Replicate** | **Conc. (pg/mL)** | **AVG conc. (pg/mL)** | **SD** | **CV%** |
| --- | --- | --- | --- | --- | --- |
| Sample 1 | Replicate 1 | 35.769 | 35.037 | 1.035 | 3% |
|  | Replicate 2 | 34.305 |  |  |  |
| Sample 2 | Replicate 1 | 58.735 | 59.156 | 0.594 | 1% |
|  | Replicate 2 | 59.576 |  |  |  |
| Sample 3 | Replicate 1 | 11.994 | 11.533 | 0.653 | 6% |
|  | Replicate 2 | 11.071 |  |  |  |
| Sample 4 | Replicate 1 | 14.300 | 13.617 | 0.965 | 7% |
|  | Replicate 2 | 12.935 |  |  |  |
| Sample 5 | Replicate 1 | 30.895 | 30.377 | 0.733 | 2% |
|  | Replicate 2 | 29.859 |  |  |  |
| Sample 6 | Replicate 1 | 4.031 | 4.033 | 0.003 | 0% |
|  | Replicate 2 | 4.035 |  |  |  |
| Sample 7 | Replicate 1 | 37.282 | 36.723 | 0.792 | 2% |
|  | Replicate 2 | 36.163 |  |  |  |
| Sample 8 | Replicate 1 | 3.373 | 3.376 | 0.004 | 0% |
|  | Replicate 2 | 3.378 |  |  |  |
| Sample 9 | Replicate 1 | 29.706 | 29.949 | 0.344 | 1% |
|  | Replicate 2 | 30.192 |  |  |  |
| Sample 10 | Replicate 1 | 2.909 | 2.952 | 0.061 | 2% |
|  | Replicate 2 | 2.996 |  |  |  |
| Sample 11 | Replicate 1 | 4.610 | 4.652 | 0.059 | 1% |
|  | Replicate 2 | 4.693 |  |  |  |
| Sample 12 | Replicate 1 | 32.567 | 33.799 | 1.742 | 5% |
|  | Replicate 2 | 35.031 |  |  |  |
| **AVG CV%** | | | | | **3%** |

**Supplementary Table 6. C231D181 assay – intra-assay variability.**

| **Cohort** | **Comparison** | **Biomarker** | **AUC** | **95% CI (Wilson/Brown)** |
| --- | --- | --- | --- | --- |
| **Discovery cohort**  **CSF** | **CSF CTRL vs. AD-dem** | p-tau181 | 1.000 | 1.000-1.000 |
|  |  | p-tau231 | 1.000 | 1.000-1.000 |
|  |  | C231D181 | 0.986 | 0.958-1.000 |
|  | **CSF CTRL vs. MCI-AD** | p-tau181 | 1.000 | 1.000-1.000 |
|  |  | p-tau231 | 1.000 | 1.000-1.000 |
|  |  | C231D181 | 0.991 | 0.968-1.000 |
| **Validation cohort**  **CSF** | **CSF CTRL vs. AD-dem** | p-tau181 | 0.997 | 0.988-1.000 |
|  |  | p-tau231 | 1.000 | 1.000-1.000 |
|  |  | C231D181 | 0.995 | 0.980-1.000 |
|  | **CSF CTRL vs. MCI-AD** | p-tau181 | 0.993 | 0.977-1.000 |
|  |  | p-tau231 | 0.991 | 0.972-1.000 |
|  |  | C231D181 | 0.995 | 0.983-1.000 |
|  | **CSF CTRL vs. pre-AD** | p-tau181 | 0.970 | 0.929-1.000 |
|  |  | p-tau231 | 0.991 | 0.972-1.000 |
|  |  | C231D181 | 0.982 | 0.951-1.000 |
|  | **CSF CTRL vs. FTD** | p-tau181 | 0.625 | 0.487-0.764 |
|  |  | p-tau231 | 0.630 | 0.491-0.769 |
|  |  | C231D181 | 0.657 | 0.521-0.793 |
|  | **CSF AD-dem vs. FTD** | p-tau181 | 0.978 | 0.940-1.000 |
|  |  | p-tau231 | 0.978 | 0.943-1.000 |
|  |  | C231D181 | 0.968 | 0.923-1.000 |
| **Discovery cohort**  **Plasma** | **Plasma CTRL vs. AD-dem** | p-tau181 | 0.943 | 0.857-1.000 |
|  |  | p-tau231 | 0.947 | 0.872-1.000 |
|  |  | C231D181 | 0.579 | 0.387-0.775 |
|  | **Plasma CTRL vs. MCI-AD** | p-tau181 | 0.877 | 0.761-0.993 |
|  |  | p-tau231 | 0.925 | 0.840-1.000 |
|  |  | C231D181 | 0.514 | 0.320-0.709 |

**Supplementary Table 7.** **Area Under Curve (AUC) values for comparison of the accuracy of each assay to discriminate between control group and different AD continuum stages (pre-AD, MCI-AD, AD-dem) in discovery and validation cohorts**. AUC values with 95% confidence intervals (CI) calculated by using Wilson/Brown method. AD: Alzheimer’s disease, pre-AD: preclinical AD, MCI-AD: mild cognitive impairment due to AD, AD-dem: AD at dementia stage, FTD: frontotemporal dementia, CTRL: control group.
